# Autonomous Screening for Diabetic Macular Edema Using Deep Learning Processing of Retinal Images

**DOI:** 10.1101/2022.08.07.22278511

**Authors:** Idan Bressler, Rachelle Aviv, Danny Margalit, Gal Yaakov Cohen, Tsontcho Ianchulev, Shravan V. Savant, David J. Ramsey, Zack Dvey-Aharon

## Abstract

**Background:** Diabetic Macular Edema (DME) is a complication of diabetes which, when untreated, leads to vision loss. Screening for signs of diabetic eye disease, including DME, is recommended for all patients with diabetes at least every one to two years, however, compliance with this standard is low.

**Methods:** A deep learning model was trained for DME detection using the EyePACS dataset. Data was randomly assigned, by participant, into development (n= 14,246) and validation (n= 1,583) sets. Analysis was conducted on the single image, eye, and patient levels. Model performance was evaluated using sensitivity, specificity, and the area under the receiver operating characteristic curve (AUC). Independent validation was further performed on the IDRiD dataset, as well as new data.

**Findings:** At the image level, sensitivity of 0.889 (CI 95% 0.878, 0.900), specificity of 0.889 (CI 95% 0.877, 0.900), and AUC of 0.954 (CI 95% 0.949, 0.959) were achieved. At the eye level, sensitivity of 0.905 (CI 95% 0.890, 0.920), specificity of 0.902 (CI 95% 0.890, 0.913), and AUC of 0.964 (CI 95% 0.958, 0.969) were achieved. At the patient level, sensitivity of 0.901 (CI 95% 0.879, 0.917), specificity of 0.900 (CI 95% 0.883, 0.911), and AUC of 0.962 (CI 95% 0.955, 0.968) were achieved.

**Interpretation:** DME can be detected from color fundus imaging with high performance on all analysis metrics. Automatic DME detection may simplify screening, leading to more encompassing screening for diabetic patients. Further prospective studies are necessary.

**Funding:** Funding was provided by AEYE Health Inc.

## Introduction

Diabetic Macular Edema (DME) is a complication of diabetes mellitus, closely associated with diabetic retinopathy (DR).^1^ DME is characterized by the accumulation of excess fluid in the extracellular space within the central macula,^2,3^ and when untreated ultimately leads to vision loss due to damage to the microvasculature and photoreceptors of the fovea, which is responsible for high-resolution visual acuity. DME has a major impact on public health, affecting approximately 3.8% of the population^4^ with an incidence of more than 25% within 25 years of diagnosis of type 1 diabetes mellitus (T1DM)^5^ and 25% within nine years of the diagnosis of type 2 diabetes mellitus (T2DM).^6^

The Early Treatment Diabetic Retinopathy Study (ETDRS) defined clinically significant macular edema (CSME) with specific anatomic criteria which include retinal thickening or the presence of hard exudates within 500 μm of the fovea.^7^ Interventions studied in the ETDRS treatment protocol, such as focal laser photocoagulation and intravitreal anti-vascular endothelial growth factor (anti-VEGF) therapy, have shown significant improvement in visual acuity and prognosis after treatment.^8–10^ Therefore, early detection and intervention are crucial in providing the opportunity to achieve good patient outcomes.

It is recommended that all patients with diabetes are screened for DME every 1 to 2 years.^11,12^ Screening is conducted as a part of regular screening and management for DR, and typically involves slit lamp examination of the dilated fundus or the use of color fundus photography.^11^ While the diagnosis of DME is traditionally based on fundus photography and fluorescein angiography, optical coherence tomography (OCT) has increasingly been used to quantitate the extent of diabetes-related retinal thickening.^13,14^ This modality is expensive, however, and generally available only at specialized eye clinics that can afford this technology; as such, it is not widely accessible for primary screening purposes. Thus, screening for DME continues to be performed by detecting classic patterns of findings in color fundus imaging, namely exudates and associated macular thickening.^10,15,16^

Access and scalability are crucial elements of any population-health screening program. The need for a specialized eye examination and sophisticated equipment have been significant obstacles for streamlined DME screening, causing many patients to remain undiagnosed.^17–20^ Machine learning and deep learning algorithms are ideal tools to address this limitation and empower an efficient and exponential screening process at the level of non-specialized, primary care delivery.^21–23^

Methods for DR screening using deep learning algorithms which examine fundus images have shown promising results.^24,25^ Furthermore, autonomous DR screening has received FDA approval,^26^ opening the door to the implementation of further similar applications, such as DME screening. While OCT-based machine learning methods have shown good results in detection of DME,^27–29^ with some methods boasting almost 100% accuracy, the cost and limited availability of OCT technology limits its ability to be used as a screening tool on a large scale.

Other fundus imagery-based methods which focus primarily on exudate detection were previously developed,^30–34^ as have been methods based on the entire CSME criteria.^35,36^ This study will expand on these works by being validated on multiple datasets, from multiple locations and using multiple definitions for CSME, showing good agreement with all of them. Furthermore, not only does the model developed exhibit excellent CSME detection, it does so at the patient level rather than by eye, making this work more clinically relevant from a functional screening standpoint.

## Methods

### Data

The main dataset utilized for training and validation was compiled and provided by EyePACS^37^ and consisted of 45° angle fundus photography images and expert readings of said images. All images and data were de-identified according to the Health Insurance Portability and Accountability Act “Safe Harbor” before they were transferred to the researchers. This study was conducted in compliance with the tenets of the Declaration of Helsinki and institutional review board exemption was obtained.

The EyePACS dataset contained up to six images per patient visit: one macula centered image, one disk centered image and one centered image per eye (in which a central fixation image is fixated on the middle of a line connecting the foveola and the optic disc). Each eye underwent expert reading, which included but was not limited to detecting the presence of DME, grading the level of DR, and assessing the image quality. It should be noted that this quality assessment was based on the overall readability of a given eye and does not guarantee that all images of an eye were of the same quality. Images deemed unreadable by an expert were omitted from our analysis, as were disc-centered images, because these provide only a limited view of the macula.

The method used to determine the presence of CSME in fundus images in the vast majority of the EyePACS dataset was Bresnick’s criterion,^38^ which is defined as the existence of hard exudates within one disc diameter from the center of the macula. A different method used in a minority of the dataset was the criterion presented in Litvin et al;^39^ this defined CSME by dividing the macula into an eight sector “pie” within one disc diameter of the center of the macula. If three sectors of the pie have hard exudates, or if hard exudates are present within 1/3 of a disc diameter from the center the macula, it was graded to have likely CSME.

A comprehensive dataset for the purposes of training, validation, and testing was constructed from the EyePACS dataset, consisting of 32,049 images from 15,892 patients. Up to two images were taken for each eye from two different fields, one centered on the macula and another centered between the macula and the disc. The average age was 55.02 (10.21 SD), and 51% of the patients were female (**Table 1**). **Table 2** shows the distribution of DME patients across DR levels; DME was only present for patients with more than mild DR (mtmDR), with approximately 49% of all images being DME positive. Additional statistics are given in **supplementary tables 1-2**

**Table 1.** Patient numbers and population statistics for the EyePACS dataset

| Field | Images | Patients | Mean Age (SD) | Gender (% Female) | Ethnicity (fraction) |
| --- | --- | --- | --- | --- | --- |
| Value | 32,049 | 15,892 | 55.02 (10.21) | 51 | White = 0.55 (Hispanic = 0.93, non-Hispanic = 0.07)<br>ethnicity not specified = 0.13<br>African Descent = 0.11<br>Indian subcontinent origin = 0.10<br>Asian = 0.03<br>Other = 0.08 |

**Table 2.**
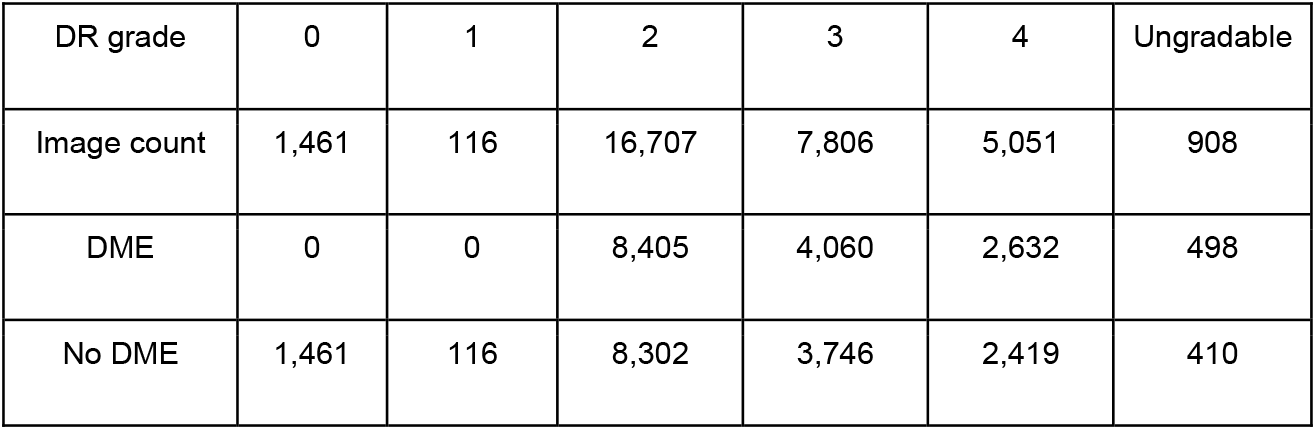
Patient number and DME prevalence across DR grades for the EyePACS dataset.

An additional dataset used for validation was the Messidor-2 dataset,^40^ also containing 45° angle fundus photography images and expert annotations for DME presence and DR level. The dataset consisted of 1748 macula-centered images from 874 examinations, of which 151 images (8.6%) were DME positive. Additional information is provided in **supplementary table 3**.

External validation was further performed on the IDRiD dataset, as well as a dataset comprised of patient images from Lahey Hospital and Medical Center.

### Pre-Processing

Image pre-processing was performed in two steps. First, the image background was cut along the convex hull which contains the circular border between the image and the background. **Figure 1** shows an example result of this process. Secondly, each image was resized to 512 × 512 pixels.

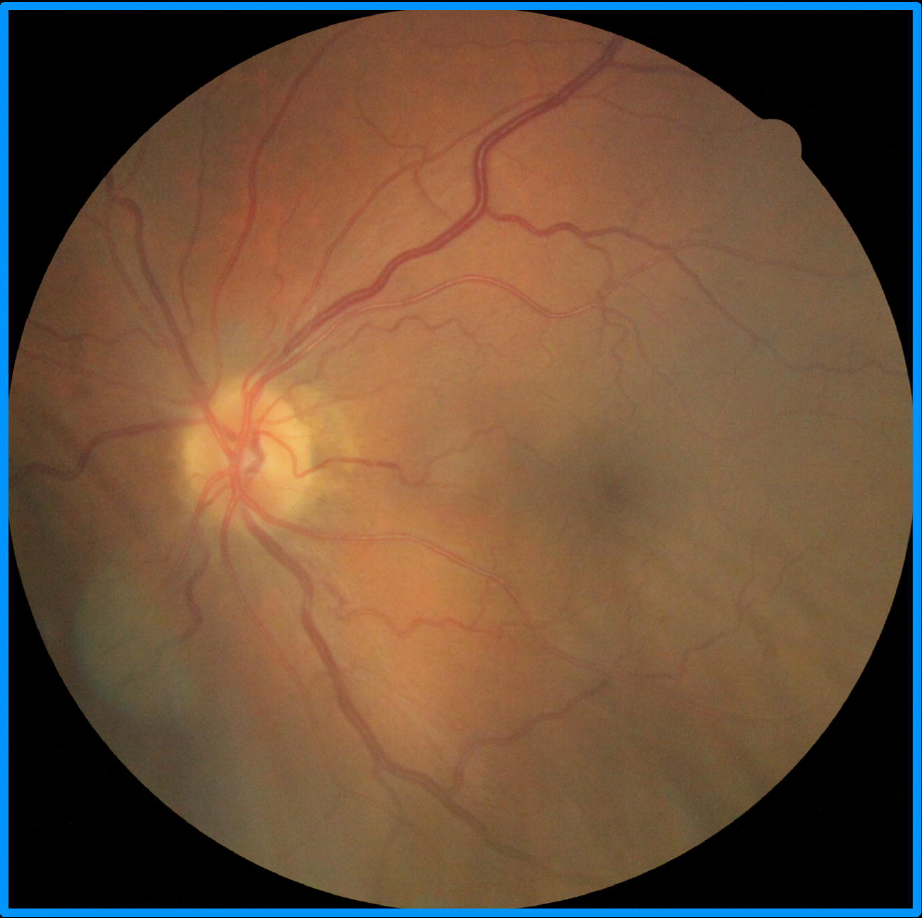

### Quality assessment

A tool for image quality assessment was developed. The tool gives a quality score for an image using an aggregation of the visibility score from multiple areas within the fundus image. **Figure 2** demonstrates a few examples of images and their respective scores, showing the correlation between score and visual image quality.

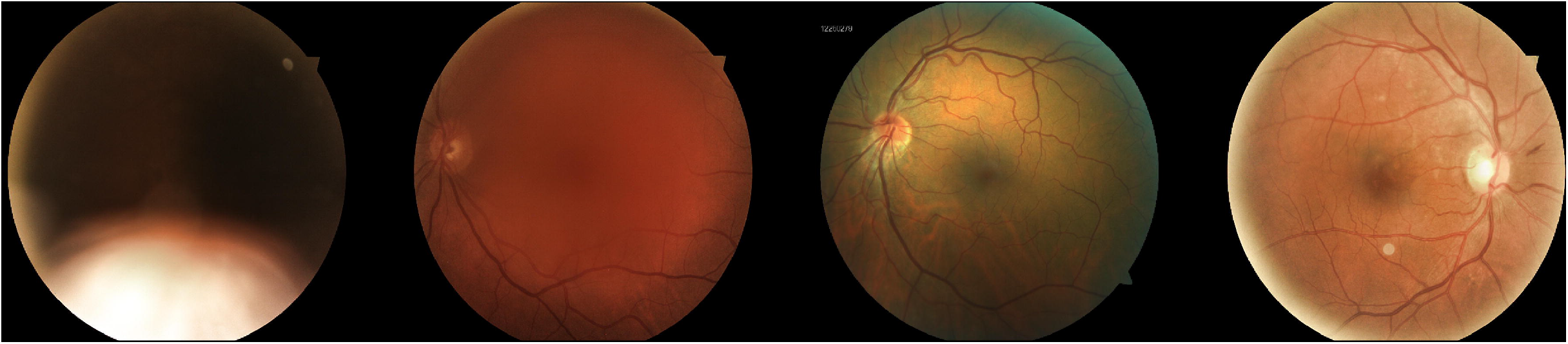

### Model training

The data was divided into training, validation, and test datasets consisting of 80%, 10%, and 10% of the data respectively.

A binary classification neural network was trained. The model architecture was automatically fitted to best balance the model performance vs model complexity tradeoff. Hyperparameter tuning was done using the validation set.

A total of 1,978 images were filtered out (approximately 6.2% of the data).

### Statistical analysis

The metrics used for assessment were accuracy, sensitivity, specificity, and area under the receiver operating characteristic curve (AUC). For each metric the bias corrected and accelerated bootstrap method^41^ was used to produce a 95% confidence interval.

### Analysis levels

DME detection was done on three different levels. The first level assessed detection at the level of each individual image, which was the basic task for which the model was trained. The second level was detection for each eye, using both macula-centered and mid-disc/macula centered image fields for a given eye. This method is akin to fundus-photo-based detection of DME performed by a human expert. In this approach the best image for each eye (in terms of image quality as assessed by the image quality tool) was selected for analysis. The third level was the patient level. For clinical purposes, detection of DME in one eye is sufficient for referral to further checks; as such, the “worst of two eyes” approached was used.

### External Validation

The module was further validated by two external teams. The first external validation set was “real-world” data collected from 50 patients with DR at the Lahey Hospital and Medical Center. Of these patients, 19 had DME confirmed by OCT and 31 did not. One macula-centered image was selected from each eye for analysis, and the evaluation was done by an ophthalmologist based on Bresnick’s method. DME detection analysis was then performed on all images using the proposed model. Performance was judged using the metrics mentioned above.

The assessment for the second external validation was conducted by an MD of The Goldschleger Eye Institute, Sheba Medical Center, Tel Hashomer, Israel, using the IDRiD training dataset,^42^ which is comprised of 400 macula-centered images. DME/CSME detection were performed based on the method presented in Wong et al,^43^ in which DME is defined via the existence of hard exudates one disc diameter from the macula and CSME is defined via the existence hard exudates 500 μm from the macula. The data was first annotated into three categories: DME positive (235), DME negative (142), and unreadable (23), and DME detection analysis was then performed on all readable images using the proposed model. Comparison was done using the metrics mentioned above.

## Results

### EyePACs dataset

The results for the different analysis methods were as follows (**table 3**), (confidence intervals set to 95% in parentheses): on the image level, sensitivity of 0.889 (0.878, 0.900) and specificity of 0.889 (0.877, 0.900) were achieved. On the eye level, sensitivity of 0.905 (0.890, 0.920) and specificity of 0.902 (0.890, 0.913) were achieved. On the patient level, sensitivity of 0.901 (0.879, 0.917) and specificity of 0.900 (0.883, 0.911) were achieved.

**Table 3.** Results for the EyePACS dataset across all three analysis levels, given in accuracy, sensitivity, specificity, and AUC with a 95% confidence interval.

|  | Accuracy (CI) | Sensitivity (CI) | Specificity (CI) | AUC (CI) |
| --- | --- | --- | --- | --- |
| Image level | 0.889 (0.881, 0.897) | 0.889 (0.878, 0.900) | 0.889 (0.877, 0.900) | 0.954 (0.949, 0.959) |
| Eye Level | 0.903 (0.894, 0.912) | 0.905 (0.890, 0.920) | 0.902 (0.890, 0.913) | 0.964 (0.958, 0.969) |
| Patient level | 0.898 (0.886, 0.909) | 0.900 (0.879, 0.917) | 0.900 (0.883, 0.911) | 0.962 (0.955, 0.968) |

The results for each DR level for which DME is present are displayed in **table 4**, showing comparable results across all DR levels. The model achieved 0.958 AUC (0.952, 0.964) for DR level 2, 0.935 AUC (0.923, 0.945) for DR level 3, 0.940 AUC (0.926, 0.952) for DR level 4, and 0.954 AUC (0.919, 0.975) for ungradable DR level. DR grades 0 and 1 did not have any DME positive examples; thus, most metrics are not defined for these grades; the model achieved an accuracy of 0.981 and 0.876 (CI not defined) respectively.

**Table 4.**
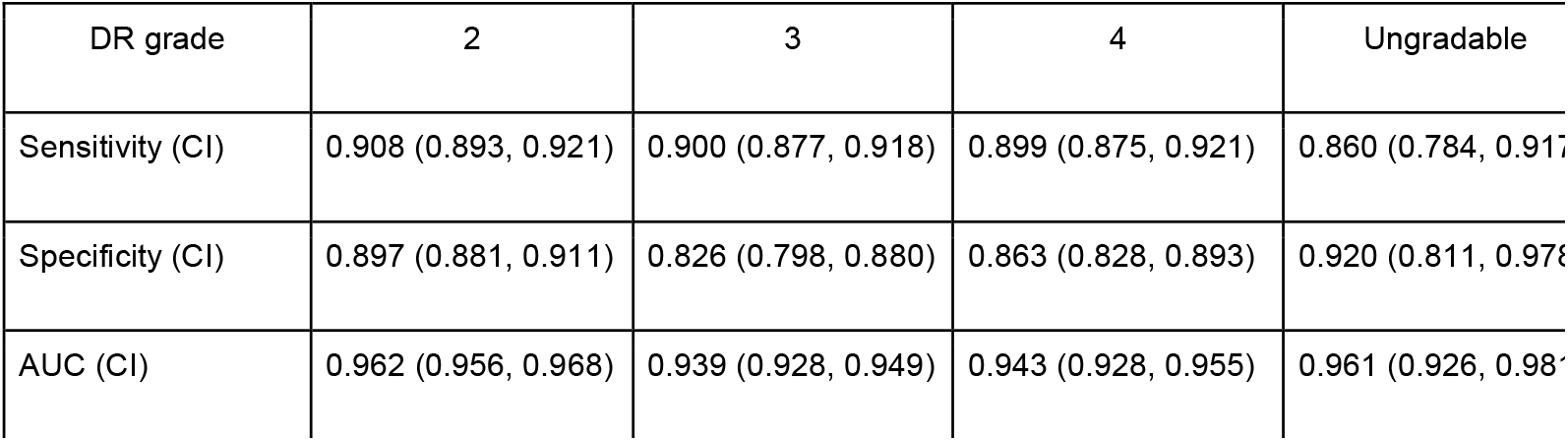
Results on the EyePACS dataset across DR grades, given in sensitivity, specificity, and AUC with a 95% confidence interval.

| DR grade | 2 | 3 | 4 | Ungradable |
| --- | --- | --- | --- | --- |
| Sensitivity (CI) | 0.908 (0.893, 0.921) | 0.900 (0.877, 0.918) | 0.899 (0.875, 0.921) | 0.860 (0.784, 0.917) |
| Specificity (CI) | 0.897 (0.881, 0.911) | 0.826 (0.798, 0.880) | 0.863 (0.828, 0.893) | 0.920 (0.811, 0.976) |
| AUC (CI) | 0.962 (0.956, 0.968) | 0.939 (0.928, 0.949) | 0.943 (0.928, 0.955) | 0.961 (0.926, 0.987) |

**Table 5** shows the results for images which passed (high quality) and didn’t pass (low quality) the quality filter, showing significant differences between the populations. The results for images which were filtered out were 0.671 sensitivity (0.599, 0.737), 0.843 specificity (0.790, 0.886), and 0.853 AUC (0.811, 0.887). Results for images which passed the quality filter were 0.902 sensitivity (0.892, 0.912), 0.883 specificity (0.871, 0.893), and 0.956 AUC (0.952, 0.961) for images that passed the filter. The filter allowed for a reading on the patient level of 98% of the patient cohort.

**Table 5.** Results for images who were filtered out and not filtered out by the image quality tool, given in accuracy, sensitivity, specificity, and AUC with a 95% confidence interval.

|  | Sensitivity (CI) | Specificity (CI) | AUC (CI) |
| --- | --- | --- | --- |
| Filtered out | 0.69 (0.611, 0.761) | 0.858 (0.805, 0.900) | 0.862 (0.819, 0.897) |
| Remained | 0.890 (0.879, 0.901) | 0.883 (0.871, 0.894) | 0.952 (0.948, 0.957) |

### Messidor-2 dataset

Messidor-2 contained readings for the image and patient levels, containing one image per eye. The results on this data set were an AUC of 0.971 (0.955 - 0.982), 0.875 sensitivity (0.811 - 0.922), and 0.954 specificity (0.939 - 0.967), surpassing previous works (**table 6**). On the patient level an AUC of 0.964 (0.936, 0.979), sensitivity of 0.897, (0.820, 0.947) and specificity of 0.932 (0.905, 0.953) were achieved.

**Table 6.** Comparison between the proposed method and previous works on the Messidor-2 dataset, given in accuracy, sensitivity, specificity, and AUC with a 95% confidence interval. Additionally, results on the patient level (not done in previous works) are given.

|  | Accuracy (CI) | Sensitivity (CI) | Specificity (CI) | AUC (CI) |
| --- | --- | --- | --- | --- |
| Sahlsten et al<sup>35</sup> | 0.931 (0.915, 0.944) | 0.69 (0.626, 0.750) | 0.989 (0.980, 0.994) | 0.932 (0.917, 0.946) |
| Li et al<sup>36</sup> | - | 0.886 (0.881, 0.892) | 0.908 (0.898, 0.912) | 0.948 (0.943, 0.951) |
| Proposed, image level | 0.943 (0.927, 0.955) | 0.875 (0.811, 0.922) | 0.954 (0.939, 0.967) | 0.971 (0.955, 0.982) |
| Proposed, patient level | 0.925 (0.898, 0.944) | 0.897 (0.820, 0.947) | 0.932 (0.905, 0.953) | 0.964 (0.936, 0.979) |

### External Validation Datasets

When tested on the first validation dataset of 100 images, the model achieved 0.880 accuracy (0.757, 0.955), 0.789 sensitivity (0.544, 0.939), and 0.935 specificity (0.786, 0.992).

When tested on the second, larger validation dataset of 413 images, 23 were labeled as unreadable. Performance of the model on the remaining 377 images demonstrated 0.854 (0.812, 0.883) accuracy, 0.851 (0.802, 0.893) sensitivity, 0.859 (0.794, 0.909) specificity and 0.931 (0.900, 0.953) AUC.

## Discussion

This work introduces a novel, proprietary autonomous system for the detection of DME from fundus images. This may shorten and simplify the screening processes and allow for wider screening of DME. Given the short (within three months) recommended referral time after DME detection,^11^ and the potential threat to patients’ vision if left untreated, the widespread use of autonomous screening has the potential to be of clinical importance.

The need to screen for DME independently from autonomous DR screening stems from three main factors. Firstly, the recommended referral time for DME is shorter than that of most DR cases without DME.^11^ Secondly, the treatment regime for DME differs from that of DR without DME,^44^ emphasizing the importance of distinguishing DME cases from DR cases. Third, the effect of DME is usually more visually significant than DR and has a higher risk of causing irreversible vision changes.^45^

This work demonstrated good results on multiple validation datasets, from multiple locations and with different definitions for CSME. This expands on previous works by showing robustness in multiple different settings and across definitions used, demonstrating the applicability and general usability of this method.

This work proposed analyzing DME on multiple levels, expanding on existing works which focused on the single image level, and shows higher efficacy on the image level as compared to previous studies. Additionally, it showed comparable results between the Messidor-2 dataset and the less curated (in terms of image quality) EyePACS dataset, demonstrating its robustness across different image qualities. The model can produce results for the vast majority of examined patients, further supporting the possible widespread capabilities and applications.

Analysis on the eye level, i.e., analyzing a single eye with multiple images of the same eye, may be more accurate and representative of clinical practice than image-level analysis. When multiple fields of the same eye exist, experts label images based on the integration of present information. This may lead to the labeling of individual images being misleading, especially if differences in image quality exist. For instance, an eye that appears healthy from one angle, often due to low image quality, might have visible DME at another angle, leading to a seemingly healthy image being positively labeled. The presented eye-level analysis tackles this issue by selecting the highest quality field from each eye.

The final model presented, which performs an analysis on the patient level, may be more clinically relevant than reporting findings at the single image or eye levels because the clinical criterion for referral is the existence of DME on the patient level. This method demonstrated high (∼90%) sensitivity and specificity.

CSME with foveal involvement is also known as CMSE with center involvement (CSME-CI),^2,11^ and the ability of graders to consistently detect this has been questioned.^46^ Despite CSME-CI being more severe, all CSME cases are referable and detection of CSME remains common clinical practice and a referral marker, thus making widespread screening of CSME important. This paper therefore focuses on CSME detection and not CSME-CI detection.

This work has a few limitations. Firstly, model training was performed on the single image level, thus hindering the training with the aforementioned image labeling problem. Secondly, the two methods used for ground truth may not be as accurate as a comprehensive eye exam using OCT in addition to the color fundus images. Finally, some of the methods used for ground truth may have underlying limitations; for example, one retrospective study examining the Bresnick method showed low specificity versus a more established ground truth (ETDRS). However, by definition, any patient with more than mild DR should be referred and most patients who have findings that meet the Bresnick criterion would qualify on this basis.

## Data Availability

Publicly available data used are available online at https://www.eyepacs.com
https://www.adcis.net/en/third-party/messidor2/
https://idrid.grand-challenge.org/

https://www.adcis.net/en/third-party/messidor2/

https://idrid.grand-challenge.org/

## Notes

### Competing Interest Statement

IB and RA are employees of AEYE Health. DM and ZDA are COO and CEO of AEYE Health.

### Funding Statement

Funding provided by AEYE Health Inc.

### Author Declarations

Sterling Institutional Review Board reviewed the study and an exemption was obtained. The Lahey Hospital institutional (Burlington, MA, USA) review board reviewed this study and an exemption was obtained.

### Summary of Updates

Additional external validation was performed on the algorithm in question and is summarized in this version of the paper.

