## supplementary table for "Autonomous Screening for Diabetic Macular Edema Using Deep Learning Processing of Retinal Images"

### Table 1:

| **Image Quality** | **Insufficient for Full Interpretation** | **Adequate** | **Good** | **Excellent** |
| --- | --- | --- | --- | --- |
| **Image Count** | 3,677 | 15,276 | 8,361 | 2,429 |
| **DME** | 1,676 | 7,658 | 4,327 | 1,424 |
| **No DME** | 2,001 | 7,618 | 4,334 | 2,005 |

Patient number and DME prevalence across annotated image quality levels for the EyePACS dataset.

### Table 2:

| **Manufacturer** | **Topcon** | **Cannon** | **Centervue** | **Crystalvue** | **Zeiss** | **Unassigned** |
| --- | --- | --- | --- | --- | --- | --- |
| **Image count** | 11,572 | 8,010 | 5,246 | 272 | 745 | 59 |
| **DME** | 3,931 | 6,135 | 3,009 | 115 | 491 | 39 |
| **No DME** | 7,641 | 14,145 | 2,237 | 167 | 254 | 20 |

Patient number and DME prevalence across camera manufacturers for the EyePACS dataset.

### Table 3:

| **DR grades** | **0** | **1** | **2** | **3** | **4** |
| --- | --- | --- | --- | --- | --- |
| **Image count** | 1017 | 270 | 347 | 75 | 35 |
| **DME** | 0 | 8 | 86 | 42 | 15 |
| **No DME** | 1017 | 262 | 261 | 33 | 20 |

Patient number and DME prevalence across DR grades for the Messidor-2 dataset
